# A Hybrid Quantum-Classical Multiscale LSTM Framework for Subject-Level EEG-Based Depression Detection

**DOI:** 10.64898/2026.05.18.26353461

**Authors:** E Sathiya, Chunzhuo Wang, T. D. Rao, T. Sunil Kumar

## Abstract

Major depressive disorder (MDD) is a common psychiatric disorder that requires reliable and objective assessment for early clinical intervention. Electroencephalography (EEG) is widely used for this purpose because it provides a non-invasive and low-cost measure of brain activity with high temporal resolution. However, EEG-based depression detection remains challenging due to the nonlinear nature of EEG signals, inter-subject variability, and the limited availability of subject-independent evaluation. To address these issues, this paper proposes a hybrid quantum–classical multiscale long short-term memory with parameterized quantum circuit branches (MS-LSTM-PQC) framework for subject-level EEG-based depression detection. The proposed model extracts temporal representations at multiple scales using parallel LSTM branches and incorporates eyes-closed (EC) and eyes-open (EO) condition information through condition-aware feature fusion. To further enhance the learned representations, scale-specific LSTM features are processed using PQC-based quantum branches implemented with TensorFlow Quantum (TFQ), providing an additional nonlinear feature transformation before classification. Experiments were conducted on the Mumtaz EEG depression dataset using EC-only, EO-only, and merged EC+EO conditions with 1-s, 2-s, and 3-s EEG windows. To reduce subject-level data leakage, all experiments were evaluated using 5-fold and 10-fold GroupKFold validation. The best overall accuracies across the evaluated settings were 92.05% and 95.08% under 5-fold and 10-fold GroupKFold validation, respectively. The 2-s merged EC+EO setting provided the most stable performance across validation protocols. In addition, Integrated Gradients (IG)-based explainability analysis showed that frontal and fronto-central channels, especially Fz, showed higher contributions to the model decision. These results suggest that multiscale temporal learning with quantum-enhanced feature transformation can support subject-level EEG-based depression detection under leakage-controlled evaluation.

## I. ntroduction

Major depressive disorder (MDD) is a common psychiatric condition that places a substantial burden on individuals and society. It affects mood, cognition, and behaviour, and in severe cases may be associated with self-harm and suicide [1]. Therefore, early and reliable identification of MDD is important for timely intervention and clinical management [2]. In current practice, diagnosis is primarily based on clinical interviews, behavioural observations, and symptom reports. Although these approaches remain essential, they depend heavily on subjective reporting and may not always capture the underlying neuro-physiological characteristics associated with depression [3]. These challenges have encouraged increasing interest in objective physiology-based methods for depression assessment.

In this context, electroencephalography (EEG) has been widely explored as a non-invasive and relatively low-cost technique for measuring brain activity with high temporal resolution [3]. However, EEG signals are nonlinear, complex, and highly variable across subjects, making manual interpretation challenging. To address this challenge, machine learning (ML) and deep learning (DL) methods have recently emerged as important tools for EEG-based classification of psychiatric disorders [3], [4].

Earlier studies on EEG-based MDD detection mainly relied on conventional ML frameworks combined with handcrafted feature extraction. For instance, the study in [3] employed traditional ML classifiers, especially support vector machine (SVM) and k-nearest neighbours (KNN), using handcrafted features from the time and frequency domains. Although such methods have shown promising results, their performance depends heavily on domain expertise for effective feature extraction [4]. In contrast, DL models can automatically learn discriminative representations directly from input data. As a result, recent research has increasingly focused on DL-based frameworks for EEG-based MDD detection. The study in [2] proposed a lightweight convolutional transformer neural network (LCTNN), which combines local temporal feature extraction with global dependency modelling while improving efficiency through adaptive channel weighting and sparse attention. Another study proposed a multi-scale multi-band attention network (MSMBANet) that captures both cross-scale temporal dependencies and cross-frequency interactions to obtain more informative temporal-spectral representations for MDD detection [5]. Similarly, the study in [6] introduced an extended one-dimensional convolutional neural network (Ex-1D-CNN) that directly learns discriminative patterns from EEG signals without manual feature engineering. In addition, an end-to-end attention-based DL model was proposed to auto-matically learn latent inter-channel connectivity patterns from resting-state EEG using multi-head self-attention, followed by a parallel convolutional module for higher-level feature extraction and classification [7].

Despite these advances, many existing DL models rely on a single representation of the signal and may not fully capture the multi-scale temporal dynamics of EEG signals [5]. In addition, although resting-state EEG is commonly acquired under eyes-closed (EC) and eyes-open (EO) conditions, such recording-condition information is not always explicitly incorporated into the learning framework [8]. Moreover, most prior studies rely exclusively on classical learning pipelines. Although quantum-based methods have been investigated in other EEG-related tasks, their application to EEG-based depression detection remains limited. To address these limitations, this study proposes a hybrid quantum–classical multi-scale LSTM framework with parameterized quantum circuit (PQC)-based branches (Hybrid MS-LSTM-PQC) for subject-level EEG-based MDD detection. The proposed model learns scale-specific temporal representations through parallel multi-scale LSTM branches, while condition information corresponding to EC and EO recordings is embedded and fused with the classical features. The learned representations are further enhanced using parallel quantum branches, and the resulting classical and quantum features are fused for final healthy control (HCs) and MDD classification.

## II. Methodology

### A. Overview of the Proposed Framework

The overall architecture is shown in Fig. 1, with the internal structure of each quantum branch. The framework is designed to learn discriminative temporal representations from resting-state EEG segments acquired under EC and EO conditions.

**Fig. 1.**
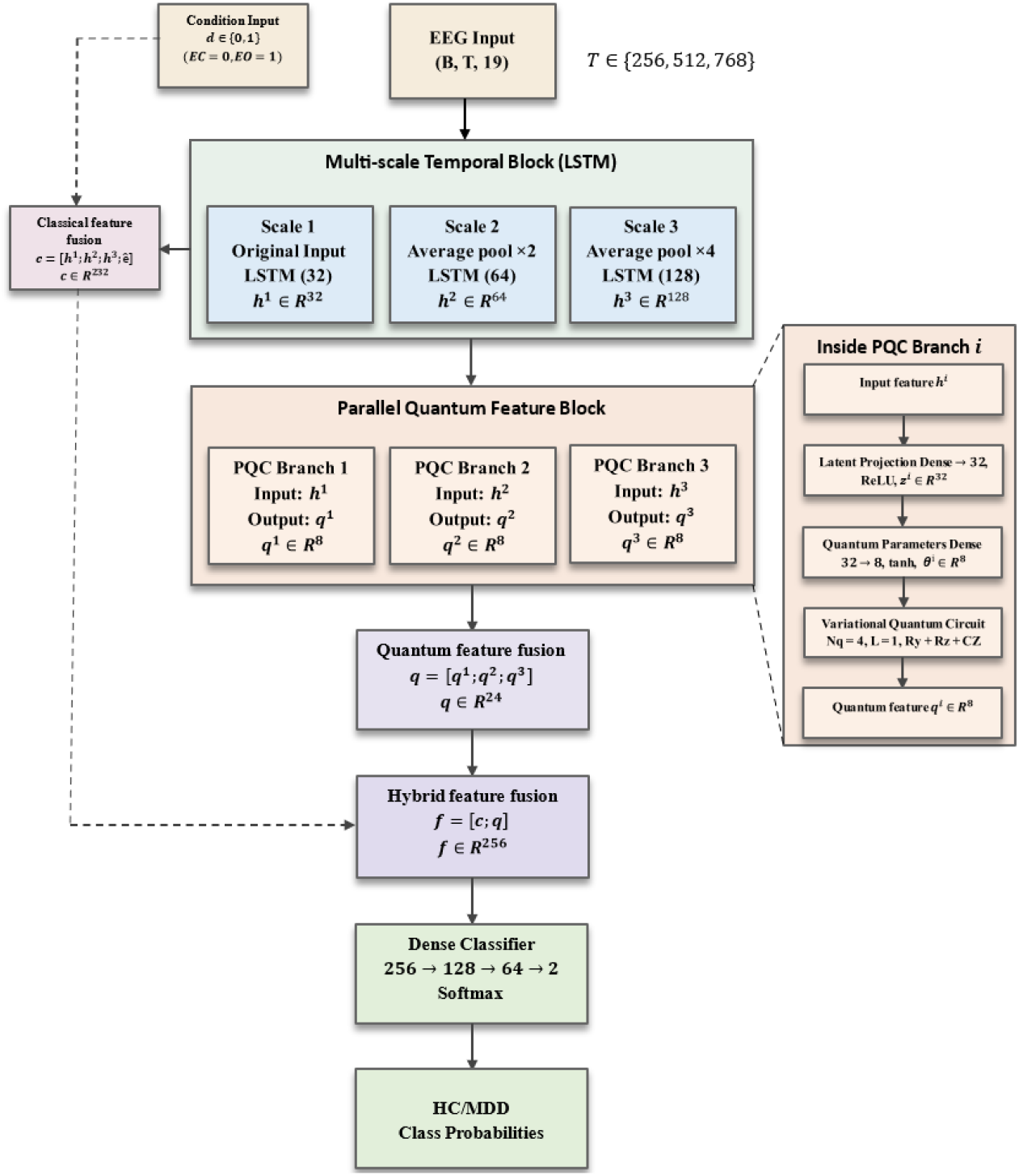
Proposed Hybrid MS-LSTM-PQC architecture. The input EEG segment is processed through three multi-scale LSTM branches, fused with the EC and EO condition feature, and further enhanced using parallel PQC branches before final HC and MDD classification.

As shown in Fig. 1, the input EEG segment is first passed through a multiscale temporal processing stage to generate three temporal representations with different resolutions. These representations are then processed by parallel LSTM branches, producing scale-specific temporal features. In addition, the EC and EO condition label is provided as an auxiliary input and incorporated into the classical feature space through a condition feature projection. For the proposed model, each scale-specific LSTM output is further passed through a dedicated quantum branch. The classical and quantum features are finally fused and fed to a dense classifier for HCs and MDD classification.

### B. Multiscale Temporal Processing

EEG signals exhibit complex temporal characteristics across multiple resolutions. To capture both fine-grained and coarse temporal dependencies, the input EEG segment is represented at three different temporal scales. Let the EEG segment be denoted as *X* ∈ ℝ^*T ×C*^, where *T* is the number of temporal samples and *C* is the number of EEG channels. Three scale-specific inputs are generated as

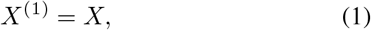

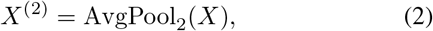

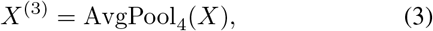

where AvgPool_2_(·) and AvgPool_4_(·) denote temporal average pooling with factors of 2 and 4, respectively. In this way, the first branch preserves the original temporal resolution, whereas the second and third branches capture progressively coarser temporal patterns [9].

Each scale-specific EEG representation is processed by an independent LSTM branch, as shown in Fig. 1. The three branches contain 32, 64, and 128 hidden units, respectively.

For the *s*-th scale, the LSTM output is represented as

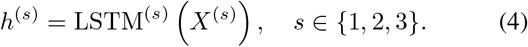

The use of separate LSTM branches enables the network to learn scale-specific temporal dependencies from the three input resolutions. Since LSTM networks are effective in modelling sequential patterns and long-range temporal dependencies [10], they are well suited for analyzing EEG segments with varying temporal characteristics.

Since the EEG recordings were acquired under EC and EO resting-state conditions, the proposed framework explicitly incorporates this recording-condition information. Let *e* ∈ {0, 1} denote the condition indicator, where 0 and 1 correspond to EC and EO, respectively. The condition feature is then fused with the multi-scale LSTM outputs to form the classical representation:

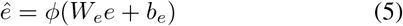

where *ϕ*(·) denotes the ReLU activation function.

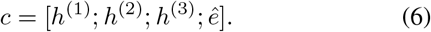

This condition-aware classical fusion enables the model to preserve both multi-scale temporal information and recording-state context.

### C. Quantum Branch Design

To further enhance the learned representations, each scale-specific LSTM output is connected to an independent quantum branch. The quantum branch receives the classical feature vector from one temporal scale and transforms it into a quantum-enhanced representation through dense projection, parameter generation, and PQC processing. PQCs are commonly used as trainable quantum models in hybrid quantum–classical frameworks [11].

More specifically, for the *i*-th scale, the scale-specific LSTM feature *h*^(*i*)^ is first passed through a dense layer to obtain a latent representation:

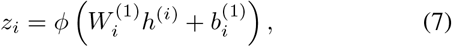

This dense projection maps the scale-specific LSTM output to a compact latent space suitable for parameter generation.

The latent vector is then mapped to a set of quantum circuit parameters:

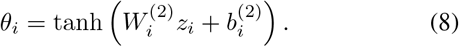

Here, *θ*_*i*_ defines the trainable rotation parameters of the quantum circuit, following the common strategy of encoding classical information into parameterized quantum rotations [11],[12]. The use of the hyperbolic tangent activation constrains the generated parameters to a bounded range before they are supplied to the circuit.

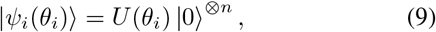

where *n* is the number of qubits. In the present work, each quantum branch uses four qubits and one variational layer. The PQC consists of parameterized single-qubit rotations followed by entangling operations. A standard representation of the circuit unitary is given by

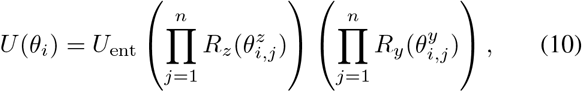

where *R*_*y*_(·) and *R*_*z*_(·) are single-qubit rotation gates and *U*_ent_ denotes the entangling block. In this work, the entangling operation is implemented using nearest-neighbour controlled-*Z* gates:

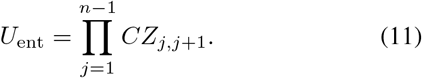

After quantum processing, the circuit produces a quantum feature vector *q*^(*i*)^ which serves as the output of the *i*-th quantum branch. Thus, for the *i*-th branch, the scale-specific LSTM feature *h*^(*i*)^ is first projected into a latent representation *z*_*i*_, then mapped to the quantum parameters *θ*_*i*_, and finally processed by the PQC *U* (*θ*_*i*_) to obtain the quantum-enhanced feature *q*^(*i*)^. This enables each temporal scale to undergo an independent quantum transformation before feature fusion. Such integration of neural-network layers with PQCs follows the hybrid quantum–classical learning approach supported by TensorFlow Quantum (TFQ) [12].

After processing the three scale-specific LSTM features through their corresponding quantum branches, the resulting quantum features are concatenated as

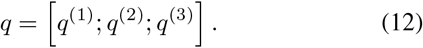

This branch-wise design allows each temporal scale to undergo an independent quantum transformation before global fusion. The resulting quantum representation is then combined with the condition-aware classical representation to form the final feature vector:

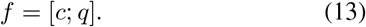

As shown in Fig. 1, the fused vector is passed through a dense classifier composed of two fully connected layers with 128 and 64 units, followed by a softmax output layer. The model is trained using subject-wise cross-validation, where subject identifiers are used as grouping labels to ensure that EEG windows from the same subject are never distributed across both training and test sets. This prevents subject-level data leakage and enables a more reliable estimate of generalization to unseen subjects.

## III. Experimental Setup and Evaluation Protocol

### A. Dataset Description

The experiments were conducted using the Mumtaz EEG depression dataset, a publicly available EEG dataset for MDD analysis [13]. The dataset contains recordings from 34 MDD subjects and 30 HCs, recorded at 256 Hz under EC and EO resting-state conditions for 5 minutes each [13], [14]. It also includes a TASK/P300 category; however, only the EC and EO recordings were used in this study because the objective here is resting-state depression detection. After preprocessing and availability checking, 58 unique subjects were available for the EC-only setting and 61 unique subjects were available for the EO-only setting. Therefore, the final usable cohort comprised 63 unique subjects (33 MDD and 30 HC). For the merged EC+EO setting, EC and EO recordings were concatenated at the window level while preserving the original subject identities. Therefore, the merged setting did not represent the sum of EC and EO subjects; instead, it contained 63 unique subjects, because subjects with both EC and EO recordings appeared under the same subject ID with different condition labels. A binary condition indicator was used to distinguish EC and EO windows during model training. In all GroupKFold experiments, grouping was performed using subject ID, ensuring that windows from the same subject, irrespective of condition, were assigned only to either the training fold or the testing fold.

Before model training, all recordings were processed using a uniform pipeline. The EDF files were first separated into EC and EO groups, followed by a 50-Hz notch filter to suppress power-line interference and a 0.5–45 Hz band-pass filter to preserve the main EEG frequency content. After filtering, each channel was standardized independently using z-score normalization. Finally, the continuous EEG recordings were segmented into fixed-length non-overlapping windows for the 1-s, 2-s, and 3-s experiments.

### B. Training Setup

To examine the effects of temporal context and recording condition, experiments were carried out using 1-s, 2-s, and 3-s EEG windows. For each window length, three data configurations were considered: EC-only, EO-only, and merged EC+EO. In the merged setting, the EC and EO window sets were concatenated while preserving subject identity, and a binary condition indicator was assigned to each segment so that the network could explicitly distinguish between the two resting states. For the EC-only and EO-only settings, the condition branch was not used, since the recording condition was fixed within each setting. Two model variants were then evaluated under each configuration: MS-LSTM and the Hybrid MS-LSTM-PQC.

Training was carried out using the Adam optimizer with a learning rate of 3 × 10^*™*4^. The loss function was sparse categorical cross-entropy, the batch size was 16, and the number of training epochs was 30. To reduce overfitting, dropout and L2 regularization were applied during training, and the random seed was fixed to 42 for reproducibility.

To avoid data leakage, GroupKFold cross-validation was used with subject identifiers as grouping labels, thereby ensuring a strictly subject-wise evaluation. Thus, all windows from a given subject were assigned either to the training fold or to the testing fold.

Two validation protocols were considered: 5-fold and 10-fold GroupKFold. Accordingly, Table I reports the results obtained under the 5-fold protocol, whereas Table II reports the results obtained under the 10-fold protocol. Under both validation settings, experiments were conducted for EC-only, EO-only, and merged EC+EO data using both the plain MS-LSTM and hybrid MS-LSTM-PQC. For subject-level evaluation, the window-level softmax probabilities of all test windows belonging to the same subject were averaged, and the class with the highest mean probability was assigned as the final subject-level prediction. For the merged EC+EO setting, this aggregation was performed across all EC and EO windows belonging to the same subject, while preserving the subject ID used for GroupKFold splitting. Although the models operate at the EEG-window level during training, the final decision is made at the subject level, since it provides a clinically more meaningful evaluation than isolated window-level classification.

**TABLE I.** Subject-level performance comparison of the proposed models under 5-fold GroupKFold for 1-s, 2-s, and 3-s eeg window lengths and recording conditions.

| Window | Model | Condition | Accuracy (%) | $\kappa$ | Precision (%) | Recall (%) | F1-score (%) |
| --- | --- | --- | --- | --- | --- | --- | --- |
| 1-s | MS-LSTM | EC+EO | 82.82 | 64.73 | 83.00 | 82.00 | 82.00 |
|  |  | EC only | 88.18 | 75.68 | 89.00 | 88.00 | 88.00 |
|  |  | EO only | 83.46 | 66.91 | 84.00 | 83.00 | 83.00 |
|  | Hybrid MS-LSTM-PQC | EC+EO | 87.43 | 74.46 | 85.71 | 90.90 | 88.23 |
|  |  | EC only | 91.66 | 82.71 | 91.00 | 91.00 | 91.00 |
|  |  | EO only | 86.79 | 73.62 | 87.00 | 87.00 | 87.00 |
| 2-s | MS-LSTM | EC+EO | 90.51 | 80.79 | 91.00 | 90.00 | 90.00 |
|  |  | EC only | 88.03 | 75.68 | 89.00 | 88.00 | 88.00 |
|  |  | EO only | 86.79 | 73.53 | 88.00 | 87.00 | 87.00 |
|  | Hybrid MS-LSTM-PQC | EC+EO | 92.05 | 84.01 | 93.00 | 92.00 | 92.00 |
|  |  | EC only | 83.03 | 65.39 | 83.00 | 83.00 | 83.00 |
|  |  | EO only | 86.79 | 73.62 | 87.00 | 87.00 | 87.00 |
| 3-s | MS-LSTM | EC+EO | 84.10 | 67.98 | 85.00 | 84.00 | 84.00 |
|  |  | EC only | 86.21 | 72.24 | 87.00 | 86.00 | 86.00 |
|  |  | EO only | 86.92 | 73.44 | 89.00 | 86.00 | 87.00 |
|  | Hybrid MS-LSTM-PQC | EC+EO | 87.30 | 74.55 | 87.88 | 87.88 | 87.88 |
|  |  | EC only | 89.84 | 79.23 | 90.00 | 90.00 | 90.00 |
|  |  | EO only | 90.12 | 80.21 | 90.00 | 90.00 | 90.00 |

**TABLE II.** Subject-level performance comparison of the proposed models under 10-fold GroupKFold for 1-s, 2-s, and 3-s eeg window lengths and recording conditions.

| Window | Model | Condition | Accuracy (%) | $\kappa$ | Precision (%) | Recall (%) | F1-score (%) |
| --- | --- | --- | --- | --- | --- | --- | --- |
| 1-s | MS-LSTM | EC+EO | 85.71 | 71.32 | 85.29 | 87.88 | 86.57 |
|  |  | EC only | 86.66 | 72.24 | 87.00 | 86.00 | 86.00 |
|  |  | EO only | 81.90 | 63.55 | 83.00 | 82.00 | 82.00 |
|  | Hybrid MS-LSTM-PQC | EC+EO | 85.71 | 71.32 | 85.29 | 87.87 | 86.56 |
|  |  | EC only | 87.93 | 75.86 | 89.66 | 86.67 | 88.14 |
|  |  | EO only | 95.08 | 90.12 | 93.94 | 96.00 | 95.38 |
| 2-s | MS-LSTM | EC+EO | 90.47 | 80.79 | 91.00 | 90.00 | 90.00 |
|  |  | EC only | 88.33 | 75.68 | 89.00 | 88.00 | 88.00 |
|  |  | EO only | 90.23 | 80.15 | 91.00 | 90.00 | 90.00 |
|  | Hybrid MS-LSTM-PQC | EC+EO | 85.47 | 71.32 | 86.00 | 86.00 | 86.00 |
|  |  | EC only | 91.66 | 82.71 | 91.00 | 91.00 | 91.00 |
|  |  | EO only | 88.57 | 76.95 | 89.00 | 88.00 | 88.00 |
| 3-s | MS-LSTM | EC+EO | 88.89 | 77.69 | 88.24 | 90.91 | 89.55 |
|  |  | EC only | 87.93 | 75.75 | 84.85 | 93.33 | 88.89 |
|  |  | EO only | 88.33 | 76.80 | 90.00 | 88.00 | 88.00 |
|  | Hybrid MS-LSTM-PQC | EC+EO | 88.81 | 77.76 | 90.62 | 87.88 | 89.23 |
|  |  | EC only | 86.66 | 72.38 | 86.00 | 86.00 | 86.00 |
|  |  | EO only | 90.25 | 80.21 | 88.23 | 93.75 | 90.90 |

### C. Performance Metrics

The classification performance was assessed at the subject level using accuracy, Cohen’s kappa (*κ*), precision, recall, and F1-score. These metrics provide complementary views of model performance and were used consistently across all experiments to compare the EC-only, EO-only, and merged EC+EO settings for both the MS-LSTM and the hybrid MS-LSTM-PQC.

## IV. Results and Discussion

### A. Results Under 5-Fold GroupKFold

The subject-level classification results obtained under the 5-fold GroupKFold protocol are shown in Table I. Across the evaluated settings, the 2-s merged EC+EO hybrid MS-LSTM-PQC achieved the best overall performance, with an accuracy of 92.05%, *κ* of 84.01%, precision of 93%, recall of 92%, and F1-score of 92%. For the EC-only setting, the highest performance was obtained by the 1-s hybrid MS-LSTM-PQC, which achieved 91.66% accuracy and 82.71% *κ*. In the EO-only setting, the best result was obtained by the 3-s hybrid MS-LSTM-PQC, which reached 90.12% accuracy and 80.21% *κ*. These results suggest that the optimal window length may vary for single-condition experiments, whereas the 2-s merged EC+EO configuration provided the strongest overall subject-level discrimination.

### B. Results Under 10-Fold GroupKFold

As shown in Table II, the highest performance was achieved by the 1-s EO-only hybrid MS-LSTM-PQC, which reached 95.08% accuracy and 90.12% *κ*. This shows that the EO condition can be highly discriminative under specific temporal settings. For the merged EC+EO configuration, however, the best result under the 10-fold protocol was obtained by the 2-s plain MS-LSTM, which achieved 90.47% accuracy and 80.79% *κ*. In the EC-only setting, the strongest performance was observed for the 2-s hybrid MS-LSTM-PQC, which achieved 91.66% accuracy. Overall, these results indicate that although the 1-s EO-only experiment gave the best individual result, the 2-s window remained the most effective choice for the merged-condition setting.

Overall, the results from both 5-fold and 10-fold GroupK-Fold evaluations show that the 2-s window length provided the most stable performance in the merged EC+EO setting. Although the highest accuracy was obtained for the 1-s EO-only quantum model under the 10-fold protocol, the 2-s merged configuration remained consistent across both validation protocols. These findings suggest that merged EC+EO analysis is effective for subject-level depression detection and that the hybrid MS-LSTM-PQC improves performance in several settings, although the gain is not uniform across all conditions.

### C. Effect of Window Length and Recording Condition

A consistent trend across both 5-fold and 10-fold subject-wise evaluation protocols is that the 2-s window length provided the most stable performance in the merged EC+EO setting. Under 5-fold evaluation, the best merged-condition result was achieved by the 2-s hybrid MS-LSTM-PQC, while under 10-fold evaluation, the best merged-condition result was achieved by the 2-s plain MS-LSTM. This indicates that the 2-s temporal duration provides a suitable balance between preserving sufficient temporal information and maintaining stable subject-level classification.

In contrast, the optimal window length for single-condition experiments was less consistent. The EC-only experiments showed good results for 1-s windows under the 5-fold protocol, whereas the EO-only setting achieved its best performance at 3-s under 5-fold and 1-s under 10-fold. These observations suggest that the temporal characteristics relevant for depression recognition may differ depending on the resting-state condition being analyzed.

### D. Comparison Between MS-LSTM and Hybrid MS-LSTM-PQC

Overall, the hybrid MS-LSTM-PQC demonstrated performance improvements in multiple experimental settings, especially under the 5-fold protocol. The most notable gain was observed in the 2-s merged EC+EO configuration, where the hybrid MS-LSTM-PQC model outperformed the MS-LSTM. Improvements were also observed for the 1-s EC-only and 3-s EO-only experiments.

However, the benefit of the quantum layer was not uniform across all configurations. In some cases, the MS-LSTM achieved comparable or better performance than the hybrid MS-LSTM-PQC variant, particularly under the 10-fold merged EC+EO setting. Therefore, the results suggest that the quantum branch can provide useful feature enhancement, but its contribution depends on the interaction between window length, recording condition, and validation protocol rather than acting as a replacement for the classical model.

### E. Comparison with the State-of-the-Art Methods

Table III compares the proposed framework with recent EEG-based depression classification methods. Existing studies have reported high accuracies using different DL models [2], [4], [7], [16]. However, these results should not be interpreted based only on accuracy values, since the studies differ in validation protocols, EEG conditions, feature representations.

**TABLE III.** Comparison with the state-of-the-art methods.

| Ref. | Dataset | Method | Validation | Accuracy (%) |
| --- | --- | --- | --- | --- |
| [4] | 33 MDD, 30 HC | 1D-CNN | 80:20 split and 10-fold cross validation | 98.32 |
| [7] | EC-only; 30 MDD, 28 HC | End-to-end DL with multi-head self-attention and parallel two-branch CNN | LOSOCV | 91.06 |
| [16] | EO and EC; 34 MDD, 30 HC | Customized Inception Time model with channel selection | Train-test split and 10-fold cross validation | 91.67 |
| [5] | EO and EC; 34 MDD, 30 HC, MODMA | MSMBANet | 80:20 train-test split | 99.70 and 99.71 |
| [18] | 34 MDD, 30 HC | MDD-AttNet | 6-fold cross validation | 99.94 |
| Our study | 33 MDD, 30 HC | MS-LSTM / Hybrid MS-LSTM-PQC | 5- and 10-GroupKFold subject-wise validation | 5-fold: 92.05;<br>10-fold: 95.08 |

Note: MDD: major depressive disorder; HC: healthy control; EC: eyes closed; EO: eyes open; DL: deep learning; CNN: convolutional neural network; LOSOCV: leave-one-subject-out cross-validation; MODMA: Multi-modal Open Dataset for Mental-disorder Analysis; MSMBANet: multi-scale multi-band attention network; MS-LSTM: multiscale long short-term memory; PQC: parameterized quantum circuit; TFQ: TensorFlow Quantum.

Recent studies [5], [18] employ multiscale and attention-based feature learning. These methods use convolutional kernels to learn local temporal patterns from EEG signals [4], [16], while MSMBANet uses multiple convolutional kernels with scale and frequency attention to capture multiscale temporal and cross-frequency information [5]. Similarly, MDD-AttNet uses multiscale convolutions, depthwise temporal modelling and attention-based refinement [18]. In comparison, the proposed Hybrid MS-LSTM-PQC framework combines multiscale temporal learning, EO and EC condition-aware fusion, and quantum-enhanced feature transformation. This allows the model to capture EEG patterns at different temporal resolutions while also considering the recording condition.

An important observation is validation protocol. Studies such as [4], [16]–[18] reported high accuracies using train-test split or conventional k-fold methods. Although these methods show strong classification performance, they may be affected by subject-level data leakage when EEG windows from the same subject are not strictly separated. In contrast, we used GroupKFold validation, where subject identity is used as the grouping factor. Therefore, all windows from one subject are placed only in either the training or testing fold. This makes the evaluation stricter and provides a reliable result for model performance on unseen subjects. Under the present GroupKFold subject-wise evaluation, the proposed framework achieved best accuracies of 92.05% and 95.08% under 5-fold and 10-fold settings, respectively, whereas [7] reported 91.06% under LOSOCV on EC-only data.

Another difference is the inclusion of recording-condition information. Many previous studies [4], [5], [7], [16], [18] evaluate either EC-only or EO-only. In the proposed study, EC-only, EO-only and merged EC+EO conditions were separately evaluated. In the merged setting, the EC and EO condition indicator was also included in the model, allowing it to distinguish between EO and EC resting-state EEG patterns. This makes the analysis more complete because the model is not limited to a single recording condition.

The proposed study also adds a Hybrid quantum–classical component through PQC-based quantum branches implemented using TFQ. While most existing methods are fully classical DL models [4], [5], [7], [16], [18], the proposed framework passes scale-specific LSTM features through quan-tum branches to provide an additional nonlinear feature transformation before final classification. The purpose of this quantum component is not to replace the classical model, but to enhance the learned multiscale temporal features. The results show that the quantum branch improved performance in several settings, although the improvement was not uniform across all window lengths and EEG conditions.

The main strengths of the proposed study are its multiscale temporal LSTM framework, EC and EO condition-aware fusion, hybrid quantum-classical feature enhancement, subject-wise validation and subject-level explainability. These make the framework more focused on unseen-subject generalization rather than only segment-level classification. However, the study also has limitations. The experiments were performed on a relatively small public dataset, and the usable subject count differed across EC, EO and merged EC+EO settings. In addition, the benefit of the quantum component was not consistent in every configuration. Hence, future work should validate the framework on larger independent datasets and further examine the stability of the quantum-enhanced feature learning.

Overall, although some existing studies report higher numerical accuracy, the proposed Hybrid MS-LSTM-PQC framework provides a more conservative and clinically meaningful comparison because it uses subject-wise validation. The study contributes by combining multiscale temporal EEG learning, recording-condition, quantum-enhanced feature transformation, and explainability for subject-level EEG-based MDD detection.

### F. Explainability Analysis

To interpret the selected 2-s merged EC+EO hybrid MS-LSTM-PQC model, integrated gradients (IG) was used to estimate channel-wise contributions to the prediction [15]. IG was computed for correctly classified test windows, and the attribution values were averaged over time across folds to obtain channel-importance scores. These scores were then summarized at global, class-wise and subject-wise levels.

The global result in Fig. 2 shows that the model assigned higher importance to frontal and fronto-central channels, especially Fz, followed by F3 and F4. This indicates that the model did not rely equally on all electrodes, but focused more on a limited set of informative channels. The class-wise analysis showed that these frontal channels contributed to both HC and MDD predictions, but with different attribution strengths. The subject-wise heatmap in Fig. 3 further shows that the frontal-dominant pattern was observed across several subjects, although inter-subject variations were present.

**Fig. 2.**
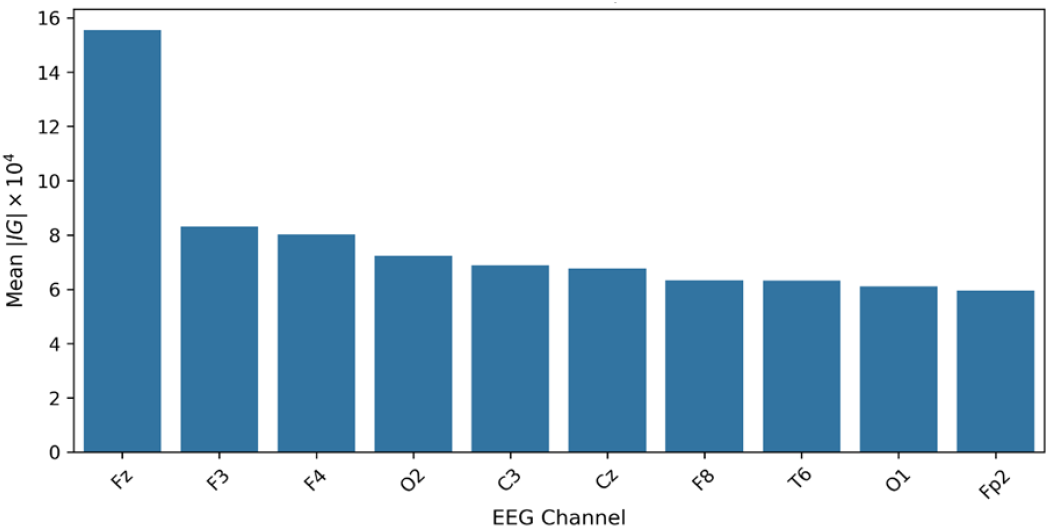
Global channel importance of the selected 2-s merged EC+EO Hybrid MS-LSTM-PQC configuration based on mean absolute IG scores.

**Fig. 3.**
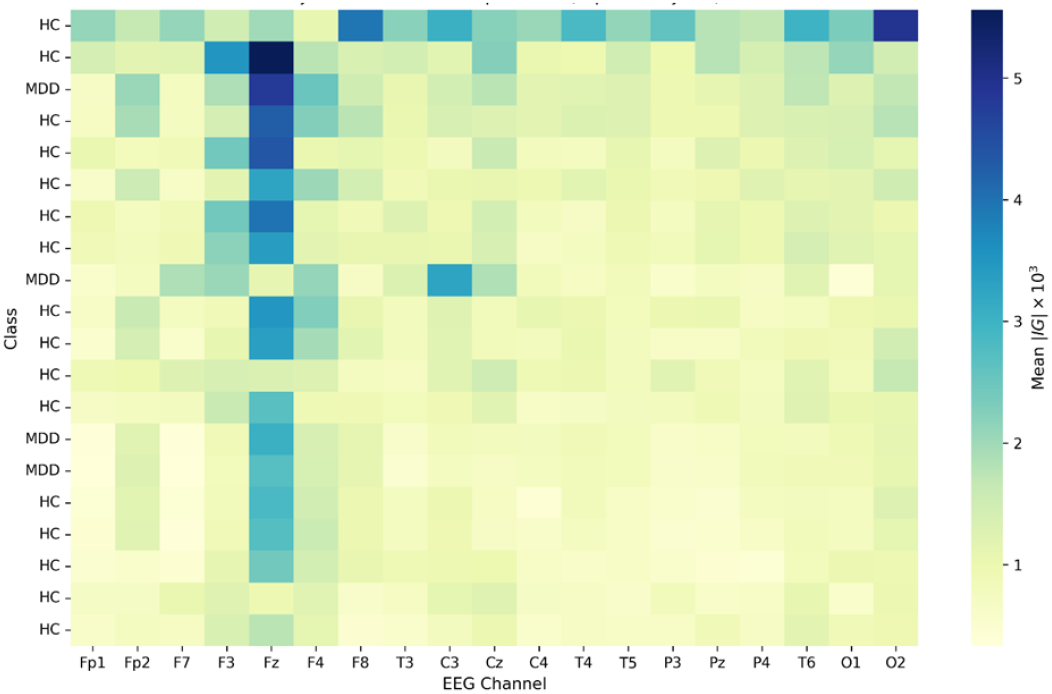
Subject-wise channel importance of the selected 2-s merged EC+EO Hybrid MS-LSTM-PQC configuration based on subject-aggregated mean absolute IG scores.

Overall, the explainability results suggest that the selected model mainly used frontal and fronto-central EEG information for subject-level MDD classification, while still showing subject-specific variation in channel contribution.

## V. Conclusion

This study proposed a hybrid quantum-classical MS-LSTM-PQC framework for subject-level EEG-based depression detection using EC-only, EO-only, and merged EC+EO EEG conditions. Experiments were performed with 1-s, 2-s and 3-s windows under 5-fold and 10-fold GroupKFold validation to avoid subject-level data leakage. The results showed that performance depended on EEG condition and window length. The 2-s merged EC+EO setting provided the most stable performance, while the best overall accuracies across the evaluated settings were 92.05% under 5-fold GroupKFold and 95.08% under 10-fold GroupKFold. The Hybrid MS-LSTM-PQC model improved performance in several settings, although the improvement was not uniform across all configurations. Overall, the findings indicate that multiscale temporal learning with condition-aware fusion and PQC-based quantum feature transformation is promising for subject-level EEG-based depression detection.

## Data Availability

The raw EEG dataset used in this study is publicly available from Figshare: "MDD patients and healthy controls EEG data (New)", DOI: 10.6084/m9.figshare.4244171.v2. The processed data generated during this study are available from the corresponding author upon reasonable request.

https://doi.org/10.6084/m9.figshare.4244171.v2

## Notes

### Competing Interest Statement

The authors have declared no competing interest.

### Funding Statement

This study did not receive any specific funding.

### Author Declarations

The study used only openly available human EEG data. The source dataset was publicly available before the initiation of this study from Figshare: "MDD patients and healthy controls EEG data (New)" by W. Mumtaz. The dataset can be located using the DOI: 10.6084/m9.figshare.4244171.v2.

